# Estimating the serial interval of the novel coronavirus disease (COVID-19): A statistical analysis using the public data in Hong Kong from January 16 to February 15, 2020

**DOI:** 10.1101/2020.02.21.20026559

**Authors:** Shi Zhao, Daozhou Gao, Zian Zhuang, Marc KC Chong, Yongli Cai, Jinjun Ran, Peihua Cao, Kai Wang, Yijun Lou, Weiming Wang, Lin Yang, Daihai He, Maggie H Wang

## Abstract

**Backgrounds:** The emerging virus, severe acute respiratory syndrome coronavirus 2 (SARS-CoV-2), has caused a large outbreak of novel coronavirus disease (COVID-19) in Wuhan, China since December 2019. Based on the publicly available surveillance data, we identified 21 transmission chains in Hong Kong and estimated the serial interval (SI) of COVID-19.

**Methods:** Index cases were identified and reported after symptoms onset, and contact tracing was conducted to collect the data of the associated secondary cases. An interval censored likelihood framework is adopted to fit a Gamma distribution function to govern the SI of COVID-19.

**Findings:** Assuming a Gamma distributed model, we estimated the mean of SI at 4.4 days (95%CI: 2.9−6.7) and SD of SI at 3.0 days (95%CI: 1.8−5.8) by using the information of all 21 transmission chains in Hong Kong.

**Conclusion:** The SI of COVID-19 may be shorter than the preliminary estimates in previous works. Given the likelihood that SI could be shorter than the incubation period, pre-symptomatic transmission may occur, and extra efforts on timely contact tracing and quarantine are recommended in combating the COVID-19 outbreak.

## Introduction

The novel coronavirus disease (COVID-19) is caused by the severe acute respiratory syndrome coronavirus 2 (SARS-CoV-2, formerly known as the ‘2019-nCoV’), which has emerged in Wuhan, China in December 2019 [1]. The COVID-19 cases were soon exported to other Chinese cities and overseas, and the travel-related risk of disease spreading was suggested by [2, 3]. Since the first confirmed imported case in Hong Kong on January 23, the local government has implemented a series of control and prevention measures for COVID-19, including enhanced border screening and traffic restrictions [4, 5].

As of February 15, there were 56 COVID-19 cases confirmed in Hong Kong [4], and local transmission was also recognized by the contact tracing investigation. Given the risk of human-to-human transmission, the serial interval (SI), which refers to the time interval from illness onset in a primary case (i.e., infector) to that in a secondary case (i.e., infectee) [6-9], was of interested to iterative rate of transmission generations of COVID-19. SI could be used to assist strategic decision-making of public health policies and construct analytical frameworks for studying the transmission dynamics of SARS-CoV-2.

In this study, we examined the publicly available materials released by the Centre for Health Protection (CHP) of Hong Kong. Adopting the case-ascertained design [10], we identified the transmission chain from index cases to secondary cases. We estimated the SI of COVID-19 based on 21 identified transmission chains from the surveillance data and contact tracing data in Hong Kong.

## Data and methods

As of February 15, there were 56 confirmed COVID-19 cases in Hong Kong [4], which followed the case definition in official diagnostic protocol released by the World Health Organization (WHO) [11]. To identify the pairs of infector (i.e., index case) and infectee (i.e., secondary case), we scanned all news press released by the CHP of Hong Kong between January 16 and February 15, 2020 [5]. The exact symptoms onset dates of all individual patients were released by CHP [4], which were publicly available, and used to match each transmission chain. For those infectees associated with multiple infectors, we record the range of onset dates of all associated infectors, i.e., lower and upper bounds. We identified 21 transmission chains, including 12 infectees matched with only one infector, that were used for SI estimation.

Following previous study [6], we adopted a Gamma distribution with mean μ and standard deviation (SD) σ, denoted by *g*(· *|μ,σ*), to govern the distribution of SI. The interval censored likelihood, denoted by *L*, of SI estimates is defined in Eqn (1).

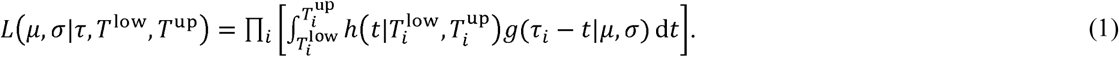

The *h*(·) was the probability density function (PDF) of exposure following a uniform distribution with a range from *T*^low^ to *T*^up^. The terms 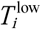 and 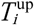 denoted the lower and upper bounds, respectively, for the range of onset dates of multiple infectors linked to the *i*-th infectee. Specially, for the infectees with only one infector, *T*^low^ = *T*^up^, and thus *h*(·) = 1. The τ_*i*_ was the observed onset date of the *i*-th infectee. We calculated the maximum likelihood estimates of *μ* and *σ*. Their 95% confidence interval (95%CI) were calculated by using the profile likelihood estimation framework with cutoff threshold determined by a Chi-square quantile [12].

In the dataset, the latest onset date of all infectors was on January 31 in contrast to the investigation period up to February 15. Given the minimum investigation period at (15 – 0 + 1 =) 16 days, which is approximately twice of the SI of the severe acute respiratory syndrome (SARS) [13], we ignored the right-truncated selection bias, i.e., ‘infector-infectee’ pairs with longer SI, due to short investigation period. Therefore, the likelihood framework defined in Eqn (1) was sufficient for the estimation in this study.

## Results and discussion

The observed SIs of all 21 samples have a mean at 4.3 days, median at 4 days, interquartile range (IQR) between 2 and 5, and range from 1 to 13 days. For the 12 ‘infector-infectee’ pairs, the observed SIs have a mean at 3 days, median at 2 days, IQR between 2 and 4, and range from 1 to 8 days. Fig 1 shows the likelihood profiles of varying SI with respect to *μ* and *σ* of SI. By using all 21 samples, we estimated the mean of SI at 4.4 days (95%CI: 2.9−6.7) and SD of SI at 3.0 days (95%CI: 1.8−5.8). The fitted Gamma distribution was shown in Fig 2. These estimates largely matched the results in the existing literatures [14, 15]. Limiting to only consider the 12 ‘infector-infectee’ pairs, we estimated the mean of SI at 3.1 days (95%CI: 2.0−5.4) and SD of SI at 1.8 days (95%CI: 1.0−4.7).

**Figure 1.**
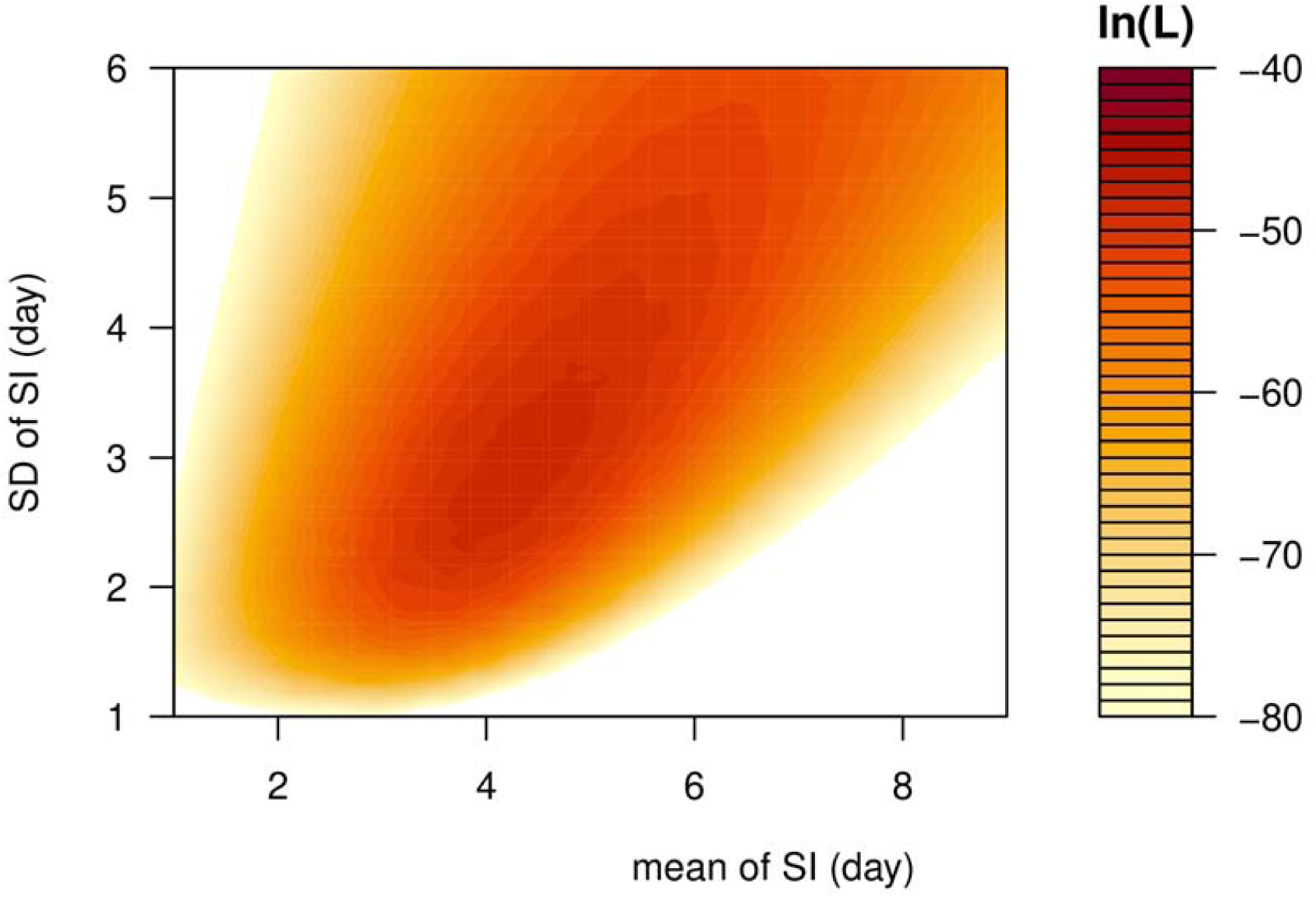
The likelihood profile of the varying serial interval (SI) of COVID-19 by using all samples. The color scheme is shown on the right-hand side, and a darker color indicates a larger log-likelihood, i.e., ln(*L*), value.

**Figure 2.**
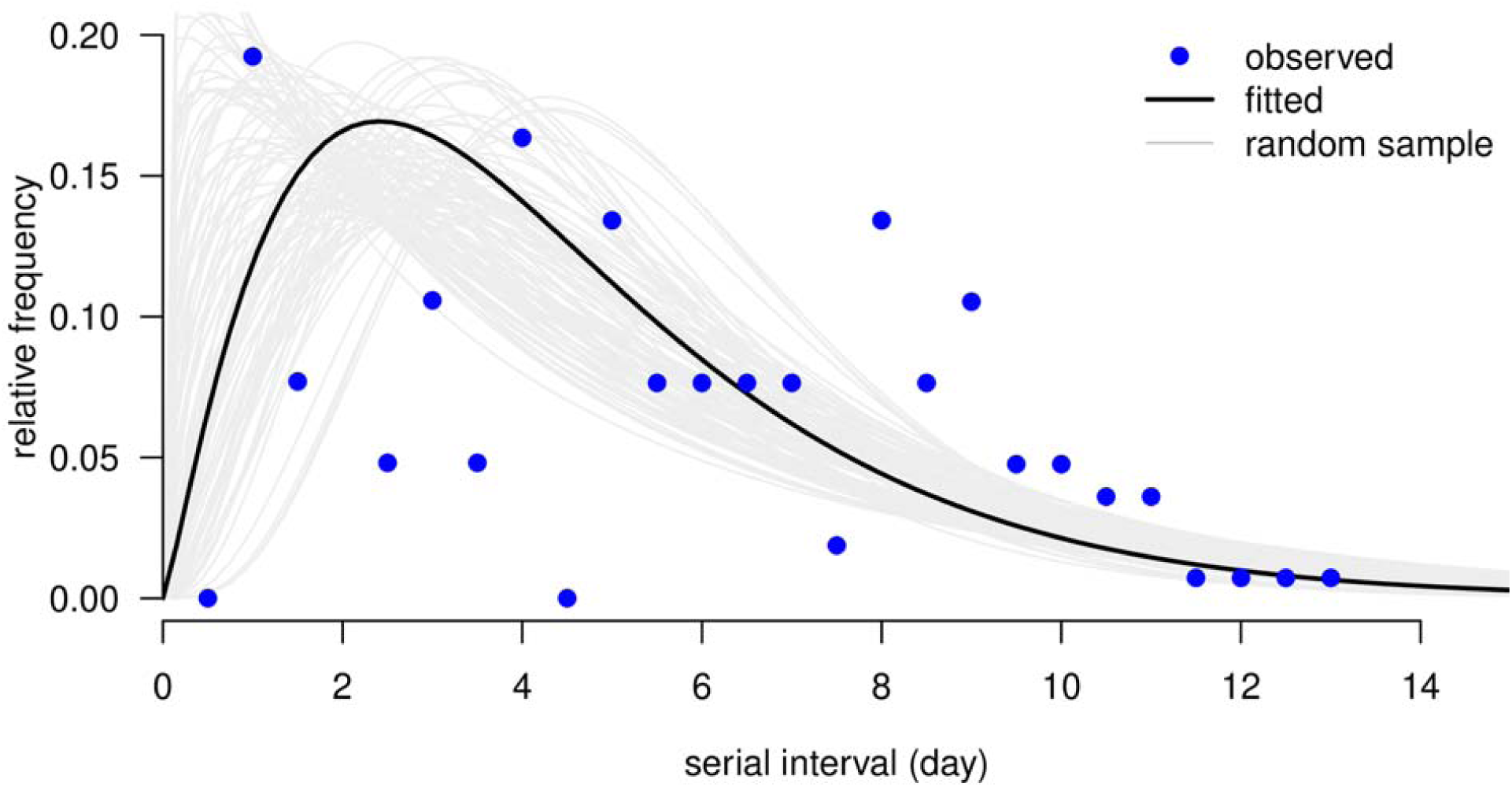
The distribution of serial interval (SI) by using all samples. The blue dots are the observed distribution of SI, the black bold curve is the fitted distribution of SI, and the light grey curves are 1000 simulation samples.

Comparing to the SI of SARS with mean at 8.4 days and SD at 3.4 days [13], the estimated 4.4-day SI for COVID-19 indicated rapid cycles of generation replacement in the transmission chain. Hence, highly efficient public health control measures, including contact tracing, isolation and screening, were strongly recommended to mitigate the epidemic size. The timely supply and delivery of healthcare resources, e.g., facemasks, alcohol sterilizer and manpower and equipment for treatment, were of required in response to the rapid growing incidences of COVID-19.

As also pointed out by two recent works [14, 15], the mean of SI at 4.4 days is notably smaller than the mean incubation period, roughly 5 days, estimated by many previous studies [16-19]. The pre-symptomatic transmission may occur when the SI is shorter than the incubation period. If isolation can be conducted immediately after the symptom onset, the pre-symptomatic transmission is likely to contribute to the most of SARS-CoV-2 infections. This situation has been recognized by a recent epidemiological investigation evidently [20], and implemented in the mechanistic modelling studies of COVID-19 epidemic [3, 21], where the pre-symptomatic cases were contagious. As such, merely isolating the symptomatic cases will lead to a considerable proportion of secondary cases, and thus contact tracing and immediately quarantine were crucial to reduce the risk of infection.

A recently epidemiological study used 5 ‘infector-infectee’ pairs from contact tracing data in Wuhan, China during the early outbreak to estimate the mean SI at 7.5 days (95%CI: 5.3−19.0) [17], which appeared larger than our SI estimate at 4.4 days. Although the 95%CIs of SI estimates in this study and [17] were not significantly separated, we note that the difference in SI might exist. If this difference was not due to sampling chance, one of the possible explanations could be the pathogenic heterogenicity in mainland China and Hong Kong. The SI estimate can be benefit from larger sample size, and the estimates in our study was backed up by 21 identified transmission chains including 12 ‘infector-infectee’ pairs, and thus it is likely to be more informative.

Accurate and consistent records on dates of illness onset were essential to the estimation of the SI. All samples used in this analysis were identified in Hong Kong and collected consistently from the CHP [4, 5]. Hence, the reporting criteria were most likely to be the same for all COVID-2019 cases, which potentially made our findings more robust.

The clusters of cases can occur by person-to-person transmission within the cluster, e.g.,

- scenario (**I**): person A infected B, C and D, or
- scenario (**II**): A to B to C to D, or
- scenario (**III**): a mixture of (**I**) and (**II**), e.g., A to B, B to C and D, or or they can occur through a common exposure to an unrecognized source of infection, e.g.,
- scenario (**IV**): unknown person X infected A, B, C and D; or
- scenario (**V**): a mixture of (**IV**) and (**I**) or (**II**), e.g., X to A and B, B to C and D; or

The lack of information in the publicly available dataset made it difficult to disentangle such complicated situations. The scenarios (**I**) and (**II**) can be covered by the pair of ‘infector-infectee’ such that we could identify the link between two unique consecutive infections. Under the scenario (**III**), we cannot clearly identify the pairwise match between the infector and infectee, which means there were multiple candidate of infector for one infectee. As such, we employed the PDF *h*(·) in Eqn (1) to account for the possible time of exposure ranging from *T*^low^ to *T*^up^. There is no information available on the SI for scenarios (**IV**) as well as (**V**) due to the onset date of person X is unknown, and thus our analysis was limited in the scenarios (**I**)-(**III**). We note that extra-cautious should be needed to interpret the clusters of cases because of this potential limitation. Although we used interval censoring likelihood to deal with the multiple-infector matching issue, more detailed information of the exposure history and clue on ‘who acquires infection from whom’ (WAIFW) would improve our estimates.

Longer SI might be difficult to occur in reality due to the isolation of confirmed infections, or to identify and link together due to the less accurate information associated with memory error occurred in the backward contact tracing exercise. The issue associated with isolation could possibly bias the SI estimates and lead to an underestimated result. Due to lack of information in the public dataset, our estimation framework could be benefit from detailed records on the date of isolation of individual cases. But isolation occurs for all severe infectious diseases such as SARS and COVID-19, there is no reason to believe that isolation play a bigger role in one than the other, at least more evidence is needed to support that claim. It is however possible that at the initial stage the SI is longer than later when strict isolation takes place. Nevertheless, a comparison of estimated SI for SARS and COVID-19 in Hong Kong is still meaningful. And we found that the SI of COVID-19 estimated appears shorter than that of SARS, which is the key message. It would be hard to imagine that isolation is responsible for the difference. There is no reason to believe isolation is more rapid in cases of COVID-19 than in SARS in Hong Kong, as well as other limitations (would have happened for both). Thus, the difference we observed for COVID-19 and SARS is likely intrinsic. In conclusion, given the rapid spreading of the COVID-19, effective contact tracing and quarantine/isolation were even more crucial for successful control.

## Data Availability

We used public available data only.

## Declarations

### Ethics approval and consent to participate

The follow-up data of individual patients were collected via public domain [4, 5], and thus neither ethical approval nor individual consent was not applicable.

### Availability of materials

All data used in this work were publicly available via [4, 5], and the exacted dataset was attached as a supplementary files of this study.

### Consent for publication

Not applicable.

## Acknowledgements

None.

## Disclaimer

The funding agencies had no role in the design and conduct of the study; collection, management, analysis, and interpretation of the data; preparation, review, or approval of the manuscript; or decision to submit the manuscript for publication.

## Authors’ Contributions

SZ conceived the study, carried out the analysis, and drafted the first manuscript. All authors discussed the results, critically read and revised the manuscript, and gave final approval for publication.

## References

1. ‘Pneumonia of unknown cause – China’, Emergencies preparedness, response, Disease outbreak news, World Health Organization (WHO) [https://www.who.int/csr/don/05-january-2020-pneumonia-of-unkown-cause-china/en/]

2. Bogoch II, Watts A, Thomas-Bachli A, Huber C, Kraemer MU, Khan K: Pneumonia of unknown etiology in Wuhan, China: potential for international spread via commercial air travel. Journal of Travel Medicine 2020:doi:10.1093/jtm/taaa1008.

3. Wu JT, Leung K, Leung GM: Nowcasting and forecasting the potential domestic and international spread of the 2019-nCoV outbreak originating in Wuhan, China: a modelling study. The Lancet 2020.

4. Summary of data and outbreak situation of the Severe Respiratory Disease associated with a Novel Infectious Agent, Centre for Health Protection, the government of Hong Kong. [https://www.chp.gov.hk/en/features/102465.html]

5. The collection of Press Releases by the Centre for Health Protection (CHP) of Hong Kong. [https://www.chp.gov.hk/en/media/116/index.html]

6. Cowling BJ, Fang VJ, Riley S, Peiris JM, Leung GM: Estimation of the serial interval of influenza. Epidemiology (Cambridge, Mass) 2009, 20(3):344.

7. Fine PEM: The Interval between Successive Cases of an Infectious Disease. American Journal of Epidemiology 2003, 158(11):1039–1047.

8. Nishiura H, Chowell G, Heesterbeek H, Wallinga J: The ideal reporting interval for an epidemic to objectively interpret the epidemiological time course. Journal of the Royal Society Interface 2010, 7(43):297–307.

9. Wallinga J, Lipsitch M: How generation intervals shape the relationship between growth rates and reproductive numbers. Proceedings of the Royal Society B: Biological Sciences 2007, 274(1609):599–604.

10. Yang Y, Longini I, Halloran ME: Design and evaluation of prophylactic interventions using infectious disease incidence data from close contact groups. J R Stat Soc Ser C-Appl Stat 2006, 55:317–330.

11. Laboratory testing for 2019 novel coronavirus (2019-nCoV) in suspected human cases, World Health Organization (WHO) [https://www.who.int/health-topics/coronavirus/laboratory-diagnostics-for-novel-coronavirus]

12. Fan J, Huang T: Profile likelihood inferences on semiparametric varying-coefficient partially linear models. Bernoulli 2005, 11(6):1031–1057.

13. Lipsitch M, Cohen T, Cooper B, Robins JM, Ma S, James L, Gopalakrishna G, Chew SK, Tan CC, Samore MH: Transmission dynamics and control of severe acute respiratory syndrome. Science 2003, 300(5627):1966–1970.

14. Nishiura H, Linton NM, Akhmetzhanov AR: Serial interval of novel coronavirus (2019-nCoV) infections. medRxiv 2020:2020.2002.2003.20019497.

15. You C, Deng Y, Hu W, Sun J, Lin Q, Zhou F, Pang CH, Zhang Y, Chen Z, Zhou X-H: Estimation of the Time-Varying Reproduction Number of COVID-19 Outbreak in China. medRxiv 2020:2020.2002.2008.20021253.

16. Backer JA, Klinkenberg D, Wallinga J: Incubation period of 2019 novel coronavirus (2019-nCoV) infections among travellers from Wuhan, China, 20–28 January 2020. Eurosurveillance 2020, 25(5):2000062.

17. Li Q, Guan X, Wu P, Wang X, Zhou L, Tong Y, Ren R, Leung KSM, Lau EHY, Wong JY et al: Early Transmission Dynamics in Wuhan, China, of Novel Coronavirus–Infected Pneumonia. New England Journal of Medicine 2020.

18. Linton MN, Kobayashi T, Yang Y, Hayashi K, Akhmetzhanov RA, Jung S-m, Yuan B, Kinoshita R, Nishiura H: Incubation Period and Other Epidemiological Characteristics of 2019 Novel Coronavirus Infections with Right Truncation: A Statistical Analysis of Publicly Available Case Data. Journal of Clinical Medicine 2020, 9(2).

19. Lauer SA, Grantz KH, Bi Q, Jones FK, Zheng Q, Meredith H, Azman AS, Reich NG, Lessler J: The incubation period of 2019-nCoV from publicly reported confirmed cases: estimation and application. medRxiv 2020:2020.2002.2002.20020016.

20. Rothe C, Schunk M, Sothmann P, Bretzel G, Froeschl G, Wallrauch C, Zimmer T, Thiel V, Janke C, Guggemos W et al: Transmission of 2019-nCoV Infection from an Asymptomatic Contact in Germany. New England Journal of Medicine 2020.

21. Chowell G, Dhillon R, Srikrishna D: Getting to zero quickly in the 2019-nCov epidemic with vaccines or rapid testing. medRxiv 2020:2020.2002.2003.20020271.

